# Coding Long COVID: Characterizing a new disease through an ICD-10 lens

**DOI:** 10.1101/2022.04.18.22273968

**Authors:** Emily R Pfaff, Charisse Madlock-Brown, John M. Baratta, Abhishek Bhatia, Hannah Davis, Andrew Girvin, Elaine Hill, Liz Kelly, Kristin Kostka, Johanna Loomba, Julie A. McMurry, Rachel Wong, Tellen D Bennett, Richard Moffitt, Christopher G Chute, Melissa Haendel, The N3C Consortium, The RECOVER Consortium

## Abstract

**Background:** Naming a newly discovered disease is a difficult process; in the context of the COVID-19 pandemic and the existence of post-acute sequelae of SARS-CoV-2 infection (PASC), which includes Long COVID, it has proven especially challenging. Disease definitions and assignment of a diagnosis code are often asynchronous and iterative. The clinical definition and our understanding of the underlying mechanisms of Long COVID are still in flux, and the deployment of an ICD-10-CM code for Long COVID in the US took nearly two years after patients had begun to describe their condition. Here we leverage the largest publicly available HIPAA-limited dataset about patients with COVID-19 in the US to examine the heterogeneity of adoption and use of U09.9, the ICD-10-CM code for “Post COVID-19 condition, unspecified.”

**Methods:** We undertook a number of analyses to characterize the N3C population with a U09.9 diagnosis code (*n* = 21,072), including assessing person-level demographics and a number of area-level social determinants of health; diagnoses commonly co-occurring with U09.9, clustered using the Louvain algorithm; and quantifying medications and procedures recorded within 60 days of U09.9 diagnosis. We stratified all analyses by age group in order to discern differing patterns of care across the lifespan.

**Results:** We established the diagnoses most commonly co-occurring with U09.9, and algorithmically clustered them into four major categories: cardiopulmonary, neurological, gastrointestinal, and comorbid conditions. Importantly, we discovered that the population of patients diagnosed with U09.9 is demographically skewed toward female, White, non-Hispanic individuals, as well as individuals living in areas with low poverty, high education, and high access to medical care. Our results also include a characterization of common procedures and medications associated with U09.9-coded patients.

**Conclusions:** This work offers insight into potential subtypes and current practice patterns around Long COVID, and speaks to the existence of disparities in the diagnosis of patients with Long COVID. This latter finding in particular requires further research and urgent remediation.

## Background

Naming diseases is an ever present challenge, and there is no shortage of efforts that aim to better standardize, disambiguate, and keep track of disease nomenclature and definitions[1–4]. Disease naming has long been controversial–for example, there are more than 400 names for syphilis dating back to the 15th century[5]. Naming a disease requires defining it, and assigning a standard code to the disease facilitates research, care, and patient engagement due to ease of patient classification and knowledge exchange. However, naming and coding a disease does not mean the disease did not exist prior to its naming or coding. For instance, although “SARS-CoV-2” and “COVID-19” were both coined February 11, 2020, by the International Committee on the Taxonomy of Viruses and the WHO, respectively[6, 7], we know that cases of COVID-19 began to surface in Wuhan, China in late December 2019[8]. In the US, most diagnostic coding uses the ICD-10-CM terminology; however the ICD-10-CM code for COVID-19, U07.1, was not made available for use until April 1, 2020. The implications of this naming delay are wide-ranging. To this day, US COVID-19 cases prior to April 1, 2020 are difficult to retrospectively ascertain. Even after that date, use of U07.1 for COVID-19 phenotyping came with caveats–use of the new code was inconsistent and of variable sensitivity and specificity, and studies have shown both underuse and overuse of U07.1 in different contexts and health systems[9–11].

Long COVID, which is included in the more general term of post-acute sequelae of SARS CoV-2 infection (PASC), is also subject to the effects of delayed naming. By Spring of 2020, patients suffering from Long COVID had coined various terms to describe the condition, including the COVID-19 long tail, long-haul COVID, and Long COVID[12–14]. Long COVID is defined by ongoing, relapsing, or new symptoms or other health effects occurring after the acute phase of SARS-CoV-2 infection (i.e., present four or more weeks after the acute infection). Heterogeneous symptoms may include, but are not limited to, fatigue, difficulty breathing, brain fog, insomnia, joint pain, and cardiac issues[15–17]. As the impact of Long COVID on health and quality of life became increasingly clear at a population level, patients worldwide came together to urge healthcare systems and policymakers to acknowledge this condition[18, 19].

Despite the relatively early recognition of this condition, an ICD-10-CM code (U09.9, “Post COVID-19 condition, unspecified”) was not made available for use in the clinical setting until October 2021. Moreover, this single code may prove insufficient: considering the phenotypic and severity variation seen in Long COVID patients, it is likely that subtypes of Long COVID exist, and such subtypes may correlate with specific underlying mechanisms that should be targeted by different interventions.

Regardless, the fact remains that there is more naming to be done, and a particular need to define and refine computable phenotypes for Long COVID and its subtypes. As can be seen by the widely differing estimates of long COVID prevalence across many studies, a lack of definitional consistency is affecting the accuracy and reproducibility of otherwise robust research.[20] Among other advantages, refined definitions will enable us to appropriately define cohorts for clinical studies, provide more precise treatment and clinical decision support, and accurately estimate long COVID’s incidence and prevalence. This is a key priority for the parent program for this work, the NIH Researching COVID to Enhance Recovery (RECOVER) Initiative,[21] which seeks to understand, treat, and prevent PASC through a wide variety of research modalities, including electronic health record (EHR) and real-world data.

In response to the COVID-19 pandemic, the US informatics and clinical community harmonized an enormous amount of EHR data to reveal candidate risk factors and therapies associated with COVID-19. The NIH’s National COVID Cohort Collaborative (N3C) is now the largest publicly available HIPAA limited EHR dataset in U.S. history, with over 14 million patients, and is a testament to the partnership of over 290 organizations. Due to the scale and demographic and geographic diversity of data within the N3C, it is uniquely well-suited to characterize the early use of the new Long COVID ICD-10-CM code. Here, we seek to characterize both (1) the early clinical use patterns of U09.9 and (2) the patients receiving that code from a provider. These characterizations reveal interesting patterns that may enable us to glean a better understanding of rough subtypes of Long COVID, current clinical practices for diagnosis and treatment of Long COVID, and potential racial and social disparities affecting who seeks and receives care for Long COVID. Ultimately, identifying patients with Long COVID based upon multiple means of inquiry (including U09.9) is critically important to recruit participants for research studies, assess the public health burden, and support nimble analytics across our heterogenous health care systems.

## Methods

To characterize the use of the U09.9 code, we used EHR data integrated and harmonized inside the NIH-hosted N3C Secure Data Enclave to identify clinical features co-occurring around the time of patients’ U09.9 index date. The methods for patient identification, data acquisition, ingestion, and harmonization into the N3C Enclave have been described previously[22–24]. Briefly, N3C contains EHR data for patients (1) who tested positive for SARS-CoV-2 infection; (2) who have a diagnosis code for COVID-19 (U07.1), multisystem inflammatory syndrome (MIS-C, M35.81), or Long COVID (U09.9); (3) whose symptoms are consistent with a COVID-19 diagnosis; or (4) are demographically matched controls who have tested negative for SARS-CoV-2 infection (and have never tested or been diagnosed as positive) to support comparative studies. Lookback data are available from January 2018 forward for each patient.

In this analysis, we defined our initial population (*n* = 23,744, sourced from 40 different health care systems) as any non-deceased patient with one or more U09.9 diagnosis codes recorded between October 1, 2021 and May 26, 2022. U09.9 codes appearing prior to October 1, 2021 may have been retroactively applied to these patients’ records (e.g., as “onset dates” in an EHR Problem List), therefore making it difficult to determine an index date that reflects the actual date of diagnosis. We excluded patients (*n* = 2,672) whose U09.9 index occurred during an inpatient hospitalization, due to the difficulty of distinguishing co-occurring clinical features related to Long COVID versus the primary reason for their hospitalization. After these exclusions, a base population of 21,072 remained. Note that we did not require patients in our cohort to have a COVID-19 diagnosis code (U07.1) or positive SARS-CoV-2 test on record, as many patients with Long COVID do not have this documentation[19].

Data from 40 of the 74 N3C sites were used for this analysis. The remaining sites either (1) did not use the U09.9 code in their N3C data or had not refreshed data since November 1, 2021, meaning the U09.9 code would not be present even if used at the site (*n =* 25 sites), or (2) did not meet the minimum criteria we set for site data for all RECOVER-related analyses (*n =* 9 sites): (a) >=25% of inpatients with at least one white blood cell count and at least one serum creatinine (to ensure lab measurement completeness); (b) 75% of inpatient visits have valid end dates; and (c) dates must not be shifted by the site more than 30 days. Additional N3C data quality criteria have been described previously, and also apply to this work.[23] The 40 sites used here are diverse in geographic location and institution size, but cannot be specifically named due to N3C governance policies.

We calculated person-level demographics and a number of social determinants of health (SDoH) variables at the area level. These variables are sourced from the Sharecare-Boston University School of Public Health Social Determinants of Health Index[25], and were linked to patients based on the preferred county (majority residence) associated with the patient’s 5-digit ZIP code. We then characterized this cohort by examining diagnoses, procedures, and medications that occurred between each patient’s U09.9 index date and 60 days after index (hereafter referred to as our “analysis window”). For each variable, values were characterized as high, medium and low based on the distribution of values across all US counties represented in the Sharecare dataset.

### Diagnosis Analysis

Our objective in characterizing diagnoses around the U09.9 index date was not only to catalog conditions and symptoms that tend to co-occur with the U09.9 diagnosis, but also to determine which of those conditions and symptoms tend to co-occur with each other. In doing so, we begin to see clusters of conditions that are more likely to occur together within a single patient’s record. First, we extracted all conditions in each patient’s record within the analysis window, and identified the most frequently occurring conditions in the study population. We then constructed an adjacency matrix for the top 30 conditions, with values indicating the frequency of co-occurrence between two conditions in the study population. From this matrix, we constructed a weighted network with nodes representing individual diagnoses, edges between nodes representing co-occurrence, and edge weights corresponding to the count of patients with both conditions. In order to detect conditions that are more likely to co-occur in our study population than at random, we tested the Louvain [26], Walktrap,[27] and Girvan-Newman[28] algorithms for community detection. We selected the Louvain algorithm in our final model, as it maximized modularity while retaining a reasonable resolution of detection. For further subgroup analyses, we present clusters detected within age-stratified condition co-occurrence networks. Additional details on community detection, network stability and subgroup analyses are available in **Supplemental Methods**.

### Procedure Analysis

Characterizing common procedures around the time of U09.9 allowed us to assess current practice patterns (i.e., diagnostics and treatments) for patients receiving the code. We defined a “procedure” as any medical diagnostics or treatments rendered by a healthcare provider. We excluded non-informative records that simply reflect that an encounter took place (e.g., CPT 99212, “Office or other outpatient visit”), despite their technical classification as “procedure codes.” We then aggregated remaining procedures into high-level categories (e.g., “radiography,” “physical therapy”) in order to discern the diagnostics and treatments that occurred within each patient’s analysis window.

### Medication Analysis

As with diagnoses and procedures, we extracted all medication records occurring within each patient’s analysis window, in order to characterize newly prescribed medications that may be used to treat symptoms of Long COVID. In order to focus on newly prescribed medications and not long-standing prescriptions, we excluded medications for each patient for which there were records prior to the patient’s U09.9 index. Medications were categorized using the third level of the Anatomical Therapeutic Chemical (ATC) classification system[29]. Results of this analysis are shown in **Supplemental Figure 2**.

## Results

Greater severity of acute SARS-CoV-2 infection does not appear to have an outsize influence in determining which patients end up with a U09.9 code; 2,542 of the U09.9 patients (12.1%) were hospitalized during a prior acute SARS-CoV-2 infection. This proportion of hospitalized patients is even lower than that cited in a recent FAIR Health white paper, which noted that 25% of patients with a U09.9 code recorded in claims data had been hospitalized with acute COVID-19.[30] Also notable is the fact that 6,806 (32.3%) of the U09.9 patients did not have a COVID index date available in N3C’s records, suggesting that these patients’ acute SARS-CoV-2 infection was indicated by a test at home, at an external health care system, or at a testing site not connected to a health system (e.g., drugstore, airport, workplace). **Table 1** shows the breakdown of the study cohort by person-level demographics and area-level social determinants of health.

**Table 1.**
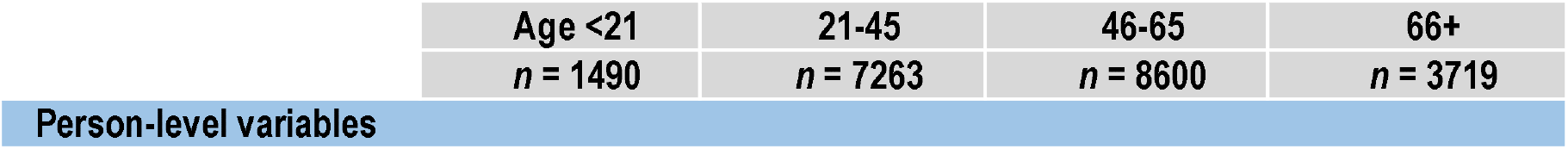
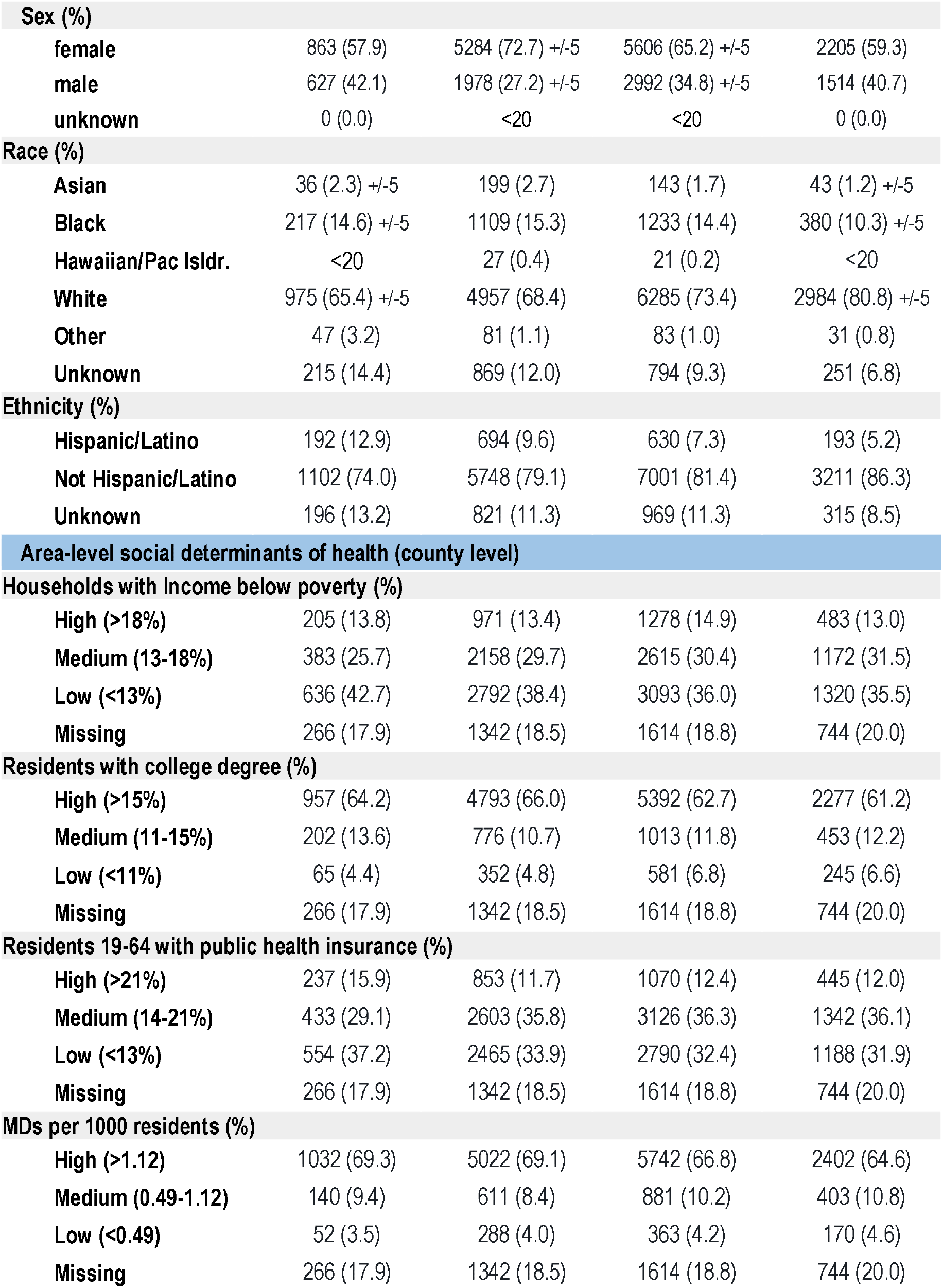
Demographic breakdown of patients in N3C with a U09.9 diagnosis code. In addition to person-level demographics, we have included a number of social determinants of health variables at the *area* level (see Methods). In accordance with the N3C download policy, for demographics where small cell sizes (<20 patients) could be derived from context, we have shifted the counts +/- by a random number between 1 and 5. The accompanying percentages reflect the shifted number. All shifted counts are labeled as such, e.g. +/- 5.

There are distinct trends among the area-level SDoH metrics. Post-hoc analysis showed that the U09.9 cohort had significantly lower representation in socially deprived counties than all COVID-19 patients in the N3C Enclave. We used the g-test of independence to compare rates in area-level SDoH across all age groups. The U09.9 patient cohort had fewer patients in the “high” category for households with income below the poverty rate compared to all COVID-19 patients (13.9% vs. 15.8%; p-value <0.01). The former cohort had a higher percentage of patients in the “high” category for residents with a college degree (63.7% vs. 51.4%; p-value <0.01), residents 19-64 with public health insurance (12.4% vs. 9.9%; p-value <0.01), and MDs per 1000 residents (67.4% vs. 60.3%; p-value <0.01).

We also analyzed uptake of the U09.9 code itself, among sites using the code. There is a rapid increase in use of U09.9 by sites following the code’s release (**Figure 1**). Usage of U09.9 post-release is compared with usage of B94.8 (“Sequelae of other specified infectious and parasitic diseases”); some sites used B94.8 at the CDC’s initial recommendation[31] as a placeholder code prior to U09.9’s release. Once U09.9 became available, it quickly supplanted B94.8.

**Figure 1.**
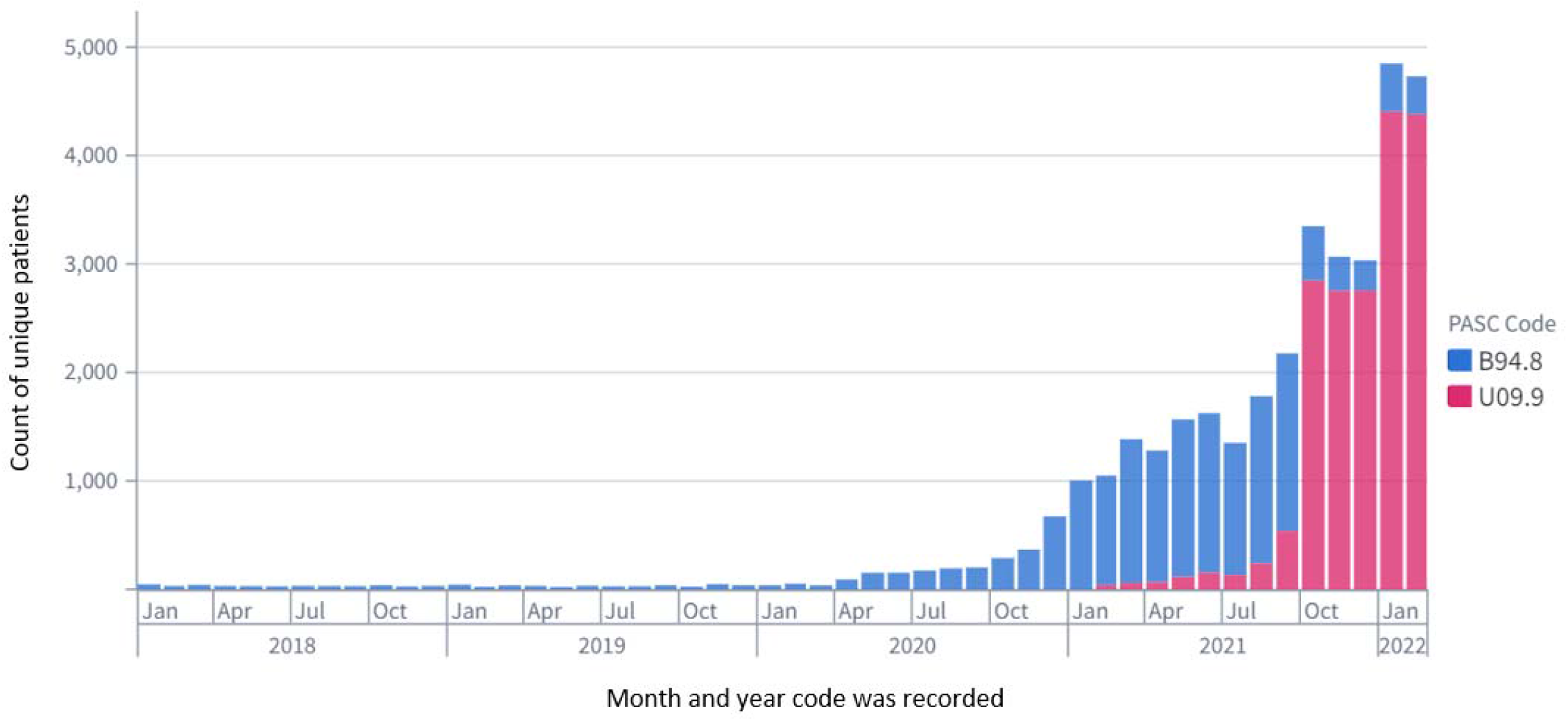
Clinical use of B94.8 decreases as U09.9 becomes available. Prior to U09.9’s release, the CDC recommended use of B94.8 (“Sequelae of other specified infectious and parasitic diseases”) as a placeholder code to signify Long COVID. As this code is not specific to sequelae of COVID-19, the figure above shows consistent but infrequent use during two pre-pandemic years. Use of B94.8 ramps up in Spring of 2020, suggesting increased recognition of Long COVID by providers. However, upon its release in October 2021, U09.9 supplants B94.8 in terms of usage frequency.

The definition of Long COVID[32] includes a wide-ranging list of symptoms and clinical features. Many of those features appear below in **Figure 2**, a visualization of diagnoses that commonly co-occur with U09.9, and each other. As shown, the mix of co-occurring diagnoses as well as the clusters produced by the Louvain algorithm change when the cohort is subset into age groups. A full accounting of diagnoses co-occurring with U09.9 (i.e., within the analysis window) in at least 20 patients from our cohort is included as **Supplemental Figure 1**.

**Figure 2.**
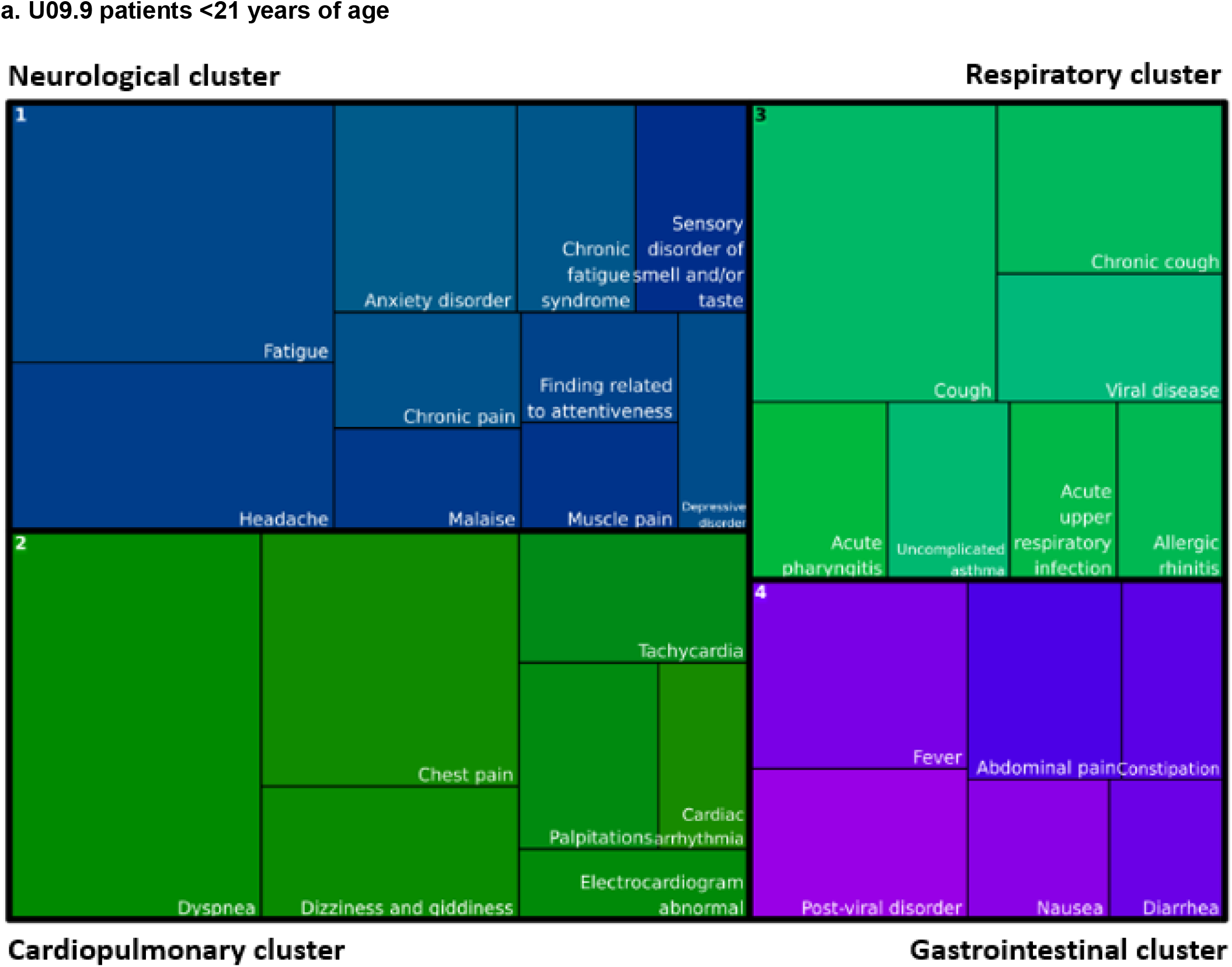
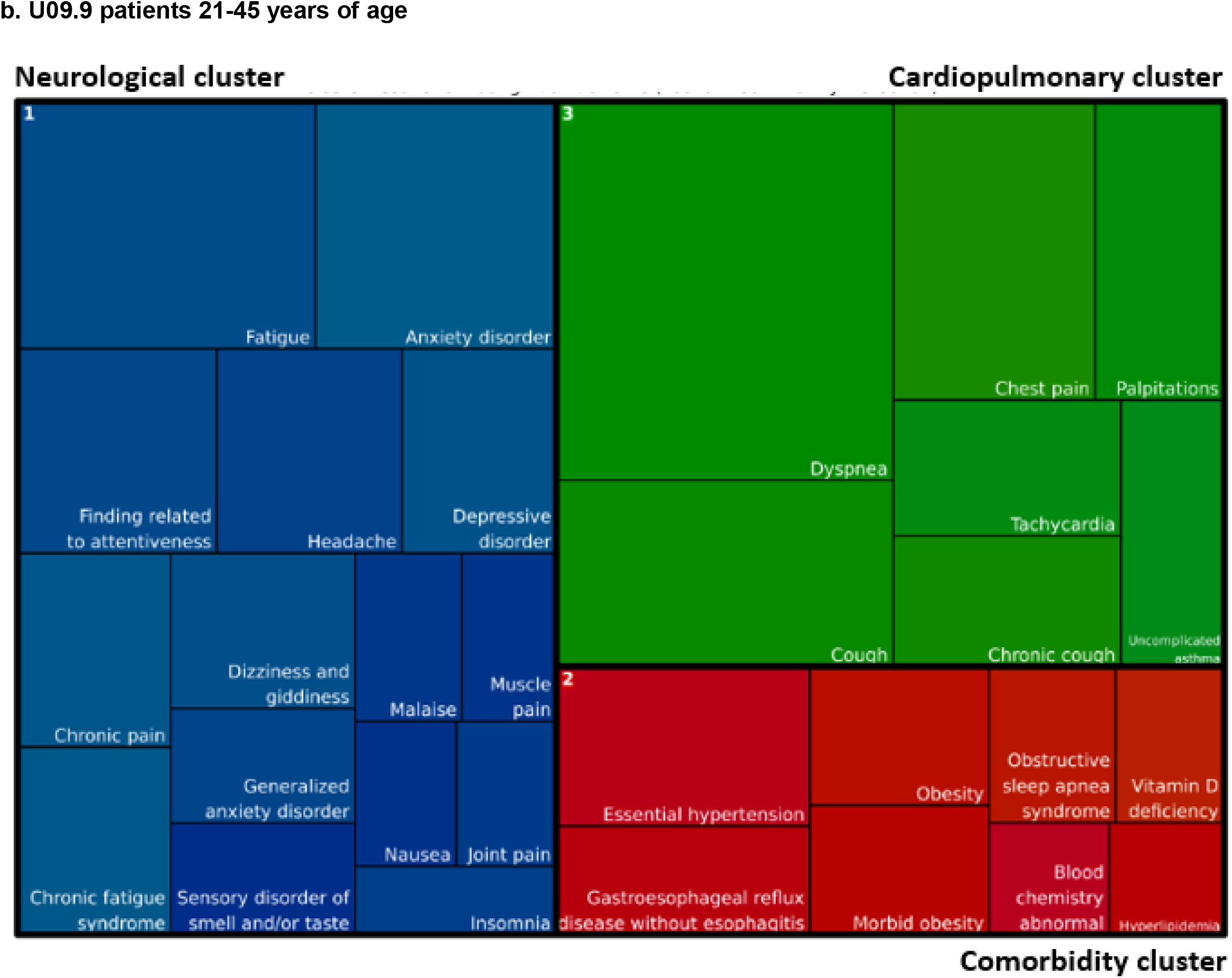
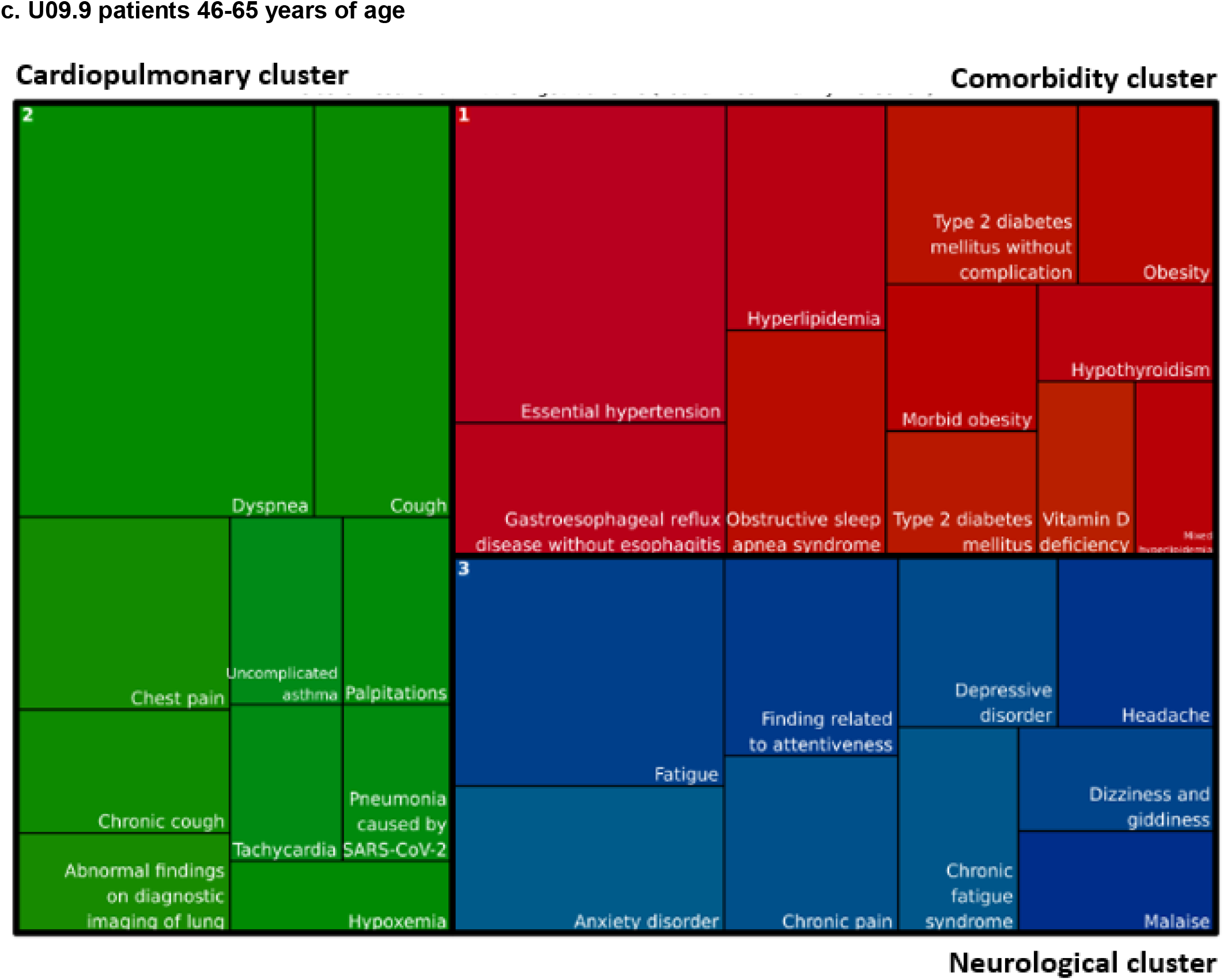
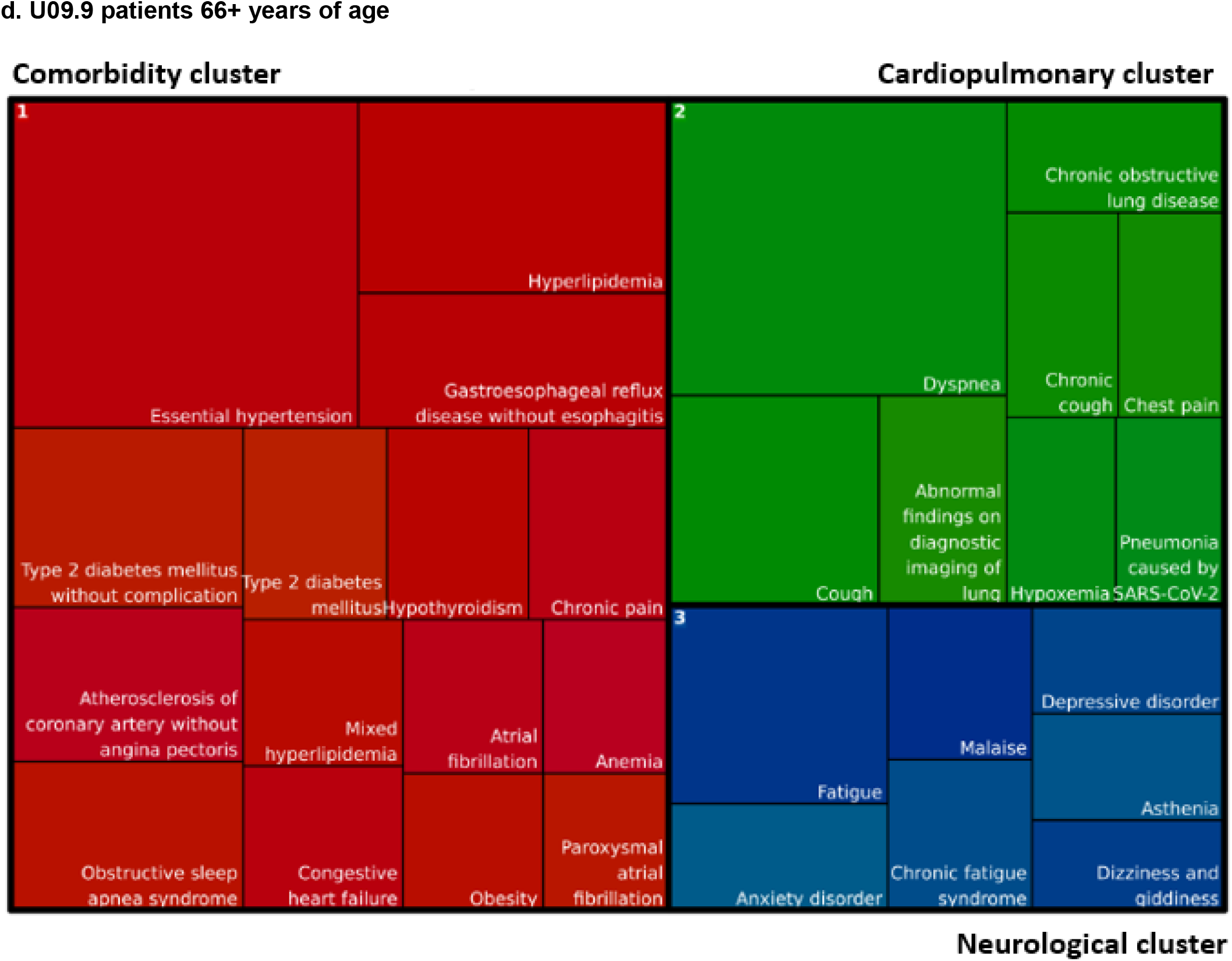
Age-stratified clusters of co-occurring diagnoses among patients with a U09.9 code. When the Louvain algorithm is applied to the top 30 most frequent pairs of co-occurring diagnoses for U09.9 patients (i.e., diagnoses co-occurring in the same patient 0 through 60 days from U09.9 diagnosis date), distinct clusters emerge. These clusters may represent rough subtypes of Long COVID presentations, and differ among age groups. The size of each box within a cluster reflects the frequency of that diagnosis relative to others in the diagram. Condition names are derived from the SNOMED CT terminology, mapped from their ICD-10-CM equivalents. Similar clusters share the same color across all four diagrams.

Our findings suggest that Long COVID symptoms and associated functional disability may present differently depending on the patient, but commonly fall into clusters. Conditions within a single cluster are more likely to co-occur within a single patient than conditions appearing in different clusters, allowing us to roughly subtype clinical presentations of Long COVID. When stratified by age, the conditions within each cluster change somewhat, though the themes remain consistent.

N3C data also enables us to examine procedures and medications that occur in each patient’s analysis window, as shown in **Figure 3** and **Supplemental Figure 2**, respectively.

**Figure 3.**
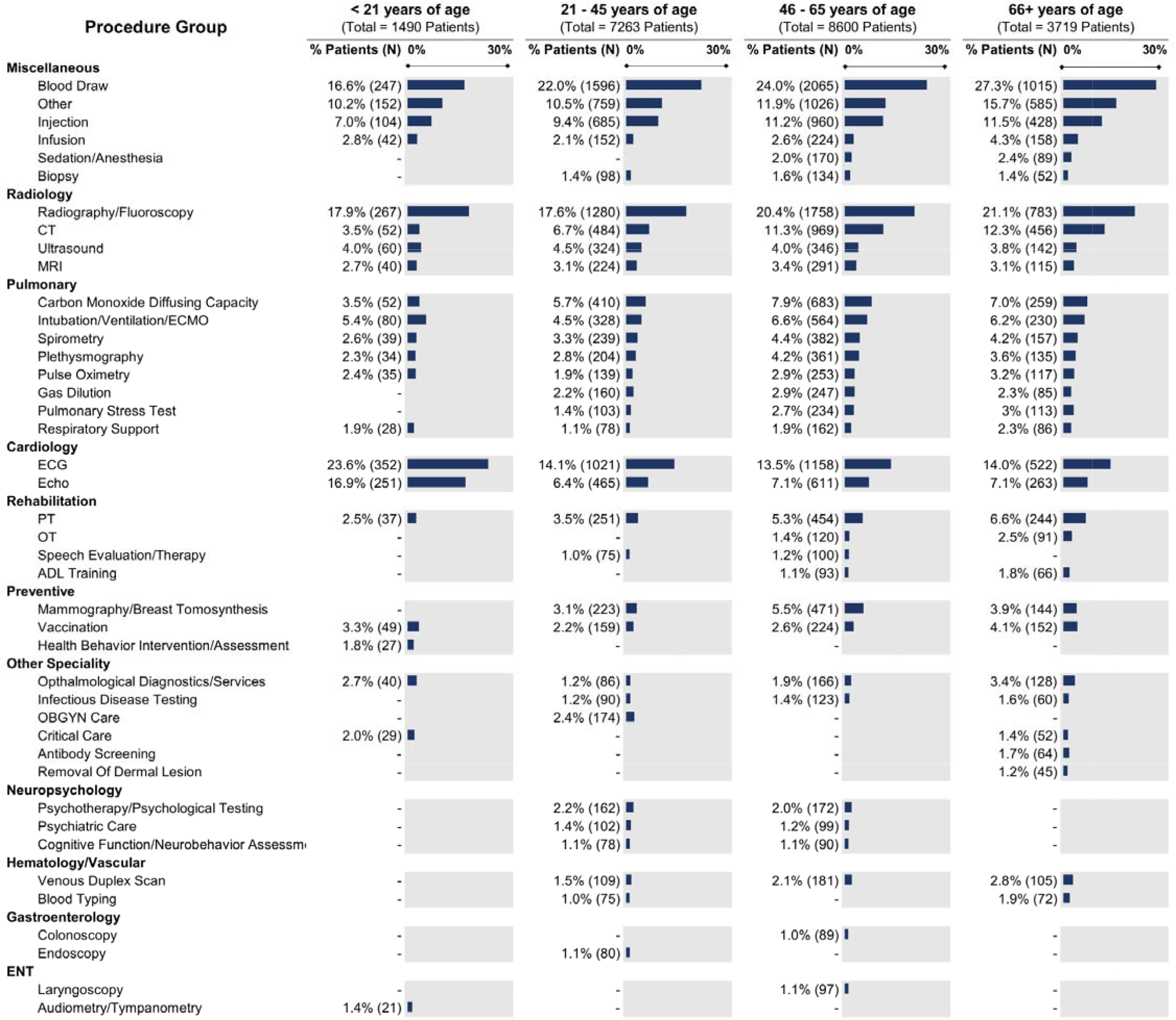
Common procedures among patients with a U09.9 code. Procedures shown occur within 60 days after a patient’s U09.9 diagnosis. Procedure records that simply reflect that an encounter took place (e.g., CPT 99212, “Office or other outpatient visit”) are excluded. Category totals represent unique patient - procedure pairs, not necessarily unique individuals. Procedure classes associated with fewer than 20 patients or less than 1.0% of the age-stratified cohort size are not shown, per N3C download policy. Percentages in each column are shown relative to the total *n* in that column.

## Discussion

Diagnosis codes are frequently used as criteria to define patient populations. While diagnosis codes alone may not define a cohort with perfect accuracy, they are a useful mechanism to narrow a population from “everyone in the EHR” to a cohort highly enriched with the condition of interest. Our analysis of U09.9 shows that this code may serve in a similar capacity to identify Long COVID patients. However, temporality and rate of uptake by providers are critical issues that must be considered. U09.9 was released for use nearly two years into the COVID-19 pandemic, resulting in potentially millions of patients with Long COVID who “missed out” on being assigned the code. Our findings must thus be interpreted through this lens of partial and incremental adoption. More work is needed to understand clinical variability and barriers to uptake by providers.

We investigated whether the use of non-specific coding such as B94.8 (“Sequelae of other specified infectious and parasitic diseases”) could be used as a proxy for early case identification. Our findings show B94.8 use increasing among COVID patients from April 2021 to October 2021, indicating a potential shift in clinical practice patterns to code for Long COVID presentation as guided by the Centers for Disease Control[31]. While B94.8 can be used for Long COVID ascertainment in EHRs prior to October 2021, it should be noted that B94.8 is used to code for any sequelae of *any* infectious disease. For this reason, it may not be specific enough to rely on for highly precise Long COVID case ascertainment without applying additional logic (e.g., requiring a positive COVID test prior to B94.8). Even still, it is likely the most reliable structured variable in the EHR to identify potential Long COVID patients prior to October 1, 2021.

Our diagnosis clusters suggest that Long COVID is not a single phenotype, but rather a collection of sub-phenotypes that may benefit from different diagnostics and treatments. Each of these clusters contains conditions and symptoms reported in existing Long COVID literature[33], clearly suggests that the definition of Long COVID is more expansive than lingering respiratory symptoms[34], and illustrates that Long COVID can manifest differently among patients in different age groups. Overall, the clusters can be summarized as neurological (in blue), cardiopulmonary (in shades of green), gastrointestinal (in purple), and comorbid conditions (in red). The clustering for the youngest patients (<21 years of age, **Figure 2a**) is the most unique, with distinct respiratory and gastrointestinal clusters that are not seen in other age groups. Patients aged 65+ (**Figure 2d**) are also unique, in that they present with more chronic diseases associated with aging (e.g. congestive heart failure, atherosclerosis, atrial fibrillation) in addition to Long COVID symptoms. The comorbid conditions cluster is unique in that it likely does not represent symptoms of Long COVID, but rather a collection of comorbid conditions that increase in incidence as patients age. The impact of these comorbid conditions on risk and outcomes of Long COVID requires further study.

Also noteworthy is the fact that the neurological cluster appears more prominently in younger groups, especially patients 21-45 years of age. Of particular note is the appearance of myalgic encephalomyelitis (listed in SNOMED CT as “chronic fatigue syndrome,” a non-preferred term)–a disease which parallels Long COVID in many ways[35–37]–in the neurological cluster across all age groups, suggesting not only frequent co-occurrence with a U09.9 diagnosis, but also co-occurrence with other neurological symptoms. The cluster differences we see among age groups make a case for age stratification when studying U09.9, and Long COVID in general. Regardless, given Long COVID’s heterogeneity in presentation, course, and outcome, the clustering of symptoms may prove informative for future development of classification and diagnostic criteria.[38]

The common procedures around the time of U09.9 index provide insight into diagnostics and treatments currently used by providers for patients presenting with Long COVID, for which treatment guidelines remain under development[39–42]. For new diseases where consensus is lacking, care is often ad hoc and informed by both the symptoms that patients present with and the available diagnostics and treatments that providers can offer. The identification and characterization of care patterns is an important step in designing future research to assess the efficacy and outcomes of these interventions. Radiographic imaging is a common occurrence across all age groups, with an average of 19.3% of patients with at least one imaging procedure in the analysis window. Electrocardiography (ECG) and echocardiography are also relatively common across all age groups, though patients younger than 21 years of age have the highest proportion (23.6% and 16.9% for ECG and echo, respectively, compared with an average of 13.9% and 6.9% across the other age groups). Pulmonary function testing shows a slight increase in frequency with more advanced age. Also of interest is the fact that some patients are receiving rehabilitation services in the 60 days after diagnosis, such as physical and occupational therapy, which lends insight into the burden of functional disability for patients with Long COVID. The proportion of patients receiving rehabilitation services also rises with patient age.

Differences across age groups were less apparent in the medication analysis (**Supplemental Figure 2**), though the youngest patients appear slightly more likely to be prescribed medications for gastrointestinal, cardiac, and neurological indications. Perhaps unsurprisingly, respiratory system drugs were the most commonly prescribed across all age groups. Interestingly, antibacterials were also used frequently across all age groups; it is unclear whether patients with Long COVID are more susceptible to bacterial infections, or if there may be overuse of antibiotics in the setting of fluctuating respiratory Long COVID symptoms or viral infections [43, 44]. Corticosteroids were also commonly used, presumably to treat persistent inflammation as a possible mechanism mediating Long COVID symptoms. The variety of medication categories seen in our analysis reflect the potential multi-system organ involvement and symptom clusters in Long COVID that we see in the analysis of conditions.

We also investigated how demographics and SDoH contribute to variation in diagnosis with U09.9. When evaluating the U09.9 cohort across age groups and SDoH variables, distinct trends can be observed (see **Table 1**). Patients with a U09.9 diagnosis code are more likely to live in counties with a high percentage of residents with college degrees and a high number of doctors per 1000 residents. Patients living in counties with a high level of poverty and/or a high percentage of residents using public insurance make up the smallest share of the U09.9 cohort. In contrast, research shows that socially deprived areas have higher rates of COVID-19 cases and deaths.[45]^,[46]^ Given the higher rates of COVID-19, lower rates of Long COVID seem unlikely. Rather, patients in deprived areas may be less likely to be treated for Long COVID. Moreover, a large majority of the U09.9 cohort identifies as female, White, and non-Hispanic. These trends are unlikely to be an accurate reflection of the true population with Long COVID, but may instead illustrate racial and social disparities in access to and experience with healthcare in the US. Clearly, the role of access to providers and the economic means to afford Long COVID care should continue to be studied for their role as contributors to disparate care and outcomes, as well as sources of research and algorithmic bias.

### Limitations

All EHR data is limited in that patients with lower access or barriers to care are less likely to be represented. EHR heterogeneity across sites may mean that a U09.9 code at one site does not quite equate to a U09.9 code at another. Moreover, we are not able to know what type of provider issued the U09.9 diagnosis (i.e., specialty), and different clinical organizations have different coding practices.

As the U09.9 code is still quite new and our sample size is limited, we cannot yet confidently label these clusters as clear “Long COVID subtypes.” Rather, these clusters are intended to be hypothesis generating, with additional work underway by the RECOVER consortium to further develop and validate these clusters. It should also be noted that many symptoms are not coded in the EHR (and may, for example, be more likely to appear in free-text notes rather than diagnosis code lists). Future work will incorporate these non-structured sources of symptoms for use in our clustering methodology.

Given the variable uptake of the U09.9 code, it is challenging to accurately identify comparator groups for this population–i.e., the absence of a U09.9 code cannot, at this time, be interpreted as the absence of Long COVID. This will continue to be an issue in future research, especially when evaluating the effect of PASC on patient morbidity and utilization of diagnostic testing and treatments.

## Conclusions

The recent release of ICD-10-CM code U09.9 to codify Long COVID will undoubtedly assist with future case ascertainment and computable phenotyping. However, a large number of patients who developed Long COVID prior to October 1, 2021 continue to be burdened with symptoms, and must also be included in data-driven cohort identification efforts for trial recruitment and retrospective analyses. Considering the caveats around rate of uptake among clinicians and late timing of the code’s release, we recommend that when characterizing Long COVID using EHRs, U09.9 should not be used alone, but rather in combination with other strategies such as more complex computable phenotypes[47]. Our findings from the characterization of patients with the U09.9 diagnosis may be of use in refining phenotypes to identify pre-U09.9 patients that might have Long COVID. There is clear utility to the characterization of early use of U09.9, as it represents the first “hook” in EHR data that can be used to identify and assess current diagnostic and treatment patterns at scale. Moreover, given the heterogeneous presentation of Long COVID, clustering of co-existing conditions and potential symptoms may be valuable in informing future development of more detailed criteria for diagnosis of Long COVID and its subtypes.

## Data Availability

The N3C data transfer to NCATS is performed under a Johns Hopkins University Reliance Protocol # IRB00249128 or individual site agreements with NIH. The N3C Data Enclave is managed under the authority of the NIH; information can be found at ncats.nih.gov/n3c/resources. Enclave data is protected, and can be accessed for COVID-related research with an approved (1) IRB protocol and (2) Data Use Request (DUR). Enclave and data access instructions can be found at https://covid.cd2h.org/for-researchers; all code used to produce the analyses in this manuscript is available within the N3C Enclave to users with valid login credentials to support reproducibility.

## Declarations

### Ethics approval

The N3C data transfer to NCATS is performed under a Johns Hopkins University Reliance Protocol # IRB00249128 or individual site agreements with NIH. The N3C Data Enclave is managed under the authority of the NIH; information can be found at https://ncats.nih.gov/n3c/resources. The work was performed under DUR RP-5677B5.

### Consent for publication

Not applicable

### Availability of data and materials

The N3C Data Enclave is managed under the authority of the NIH; information can be found at ncats.nih.gov/n3c/resources. Enclave data is protected, and can be accessed for COVID-related research with an approved (1) IRB protocol and (2) Data Use Request (DUR). A detailed accounting of data protections and access tiers is found in [1]. Enclave and data access instructions can be found at https://covid.cd2h.org/for-researchers; all code used to produce the analyses in this manuscript is available within the N3C Enclave to users with valid login credentials to support reproducibility.

### Competing interests

Author ATG is an employee of Palantir Technologies. MAH and JAM are co-founders of Pryzm Health.

### Funding

This research was funded by the National Institutes of Health (NIH) Agreement OT2HL161847-01. The views and conclusions contained in this document are those of the authors and should not be interpreted as representing the official policies, either expressed or implied, of the NIH.

The analyses described in this publication were conducted with data or tools accessed through the NCATS N3C Data Enclave covid.cd2h.org/enclave and supported by NCATS U24 TR002306.

#### Data Partners with Released Data

Stony Brook University — U24TR002306 • University of Oklahoma Health Sciences Center — U54GM104938: Oklahoma Clinical and Translational Science Institute (OCTSI) • West Virginia University — U54GM104942: West Virginia Clinical and Translational Science Institute (WVCTSI) • University of Mississippi Medical Center U54GM115428: Mississippi Center for Clinical and Translational Research (CCTR) • University of Nebraska Medical Center — U54GM115458: Great Plains IDeA-Clinical & Translational Research • Maine Medical Center — U54GM115516: Northern New England Clinical & Translational Research (NNE-CTR) Network • Wake Forest University Health Sciences — UL1TR001420: Wake Forest Clinical and Translational Science Institute • Northwestern University at Chicago — UL1TR001422: Northwestern University Clinical and Translational Science Institute (NUCATS) • University of Cincinnati — UL1TR001425: Center for Clinical and Translational Science and Training • The University of Texas Medical Branch at Galveston — UL1TR001439: The Institute for Translational Sciences • Medical University of South Carolina — UL1TR001450: South Carolina Clinical & Translational Research Institute (SCTR) • University of Massachusetts Medical School Worcester — UL1TR001453: The UMass Center for Clinical and Translational Science (UMCCTS) • University of Southern California — UL1TR001855: The Southern California Clinical and Translational Science Institute (SC CTSI) • Columbia University Irving Medical Center — UL1TR001873: Irving Institute for Clinical and Translational Research • George Washington Children’s Research Institute — UL1TR001876: Clinical and Translational Science Institute at Children’s National (CTSA-CN) • University of Kentucky — UL1TR001998: UK Center for Clinical and Translational Science • University of Rochester — UL1TR002001: UR Clinical & Translational Science Institute • University of Illinois at Chicago — UL1TR002003: UIC Center for Clinical and Translational Science • Penn State Health Milton S. Hershey Medical Center — UL1TR002014: Penn State Clinical and Translational Science Institute • The University of Michigan at Ann Arbor — UL1TR002240: Michigan Institute for Clinical and Health Research • Vanderbilt University Medical Center — UL1TR002243: Vanderbilt Institute for Clinical and Translational Research • University of Washington — UL1TR002319: Institute of Translational Health Sciences • Washington University in St. Louis — UL1TR002345: Institute of Clinical and Translational Sciences • Oregon Health & Science University — UL1TR002369: Oregon Clinical and Translational Research Institute • University of Wisconsin-Madison — UL1TR002373: UW Institute for Clinical and Translational Research • Rush University Medical Center — UL1TR002389: The Institute for Translational Medicine (ITM) • The University of Chicago — UL1TR002389: The Institute for Translational Medicine (ITM) • University of North Carolina at Chapel Hill — UL1TR002489: North Carolina Translational and Clinical Science Institute • University of Minnesota — UL1TR002494: Clinical and Translational Science Institute • Children’s Hospital Colorado — UL1TR002535: Colorado Clinical and Translational Sciences Institute • The University of Iowa — UL1TR002537: Institute for Clinical and Translational Science • The University of Utah — UL1TR002538: Uhealth Center for Clinical and Translational Science • Tufts Medical Center — UL1TR002544: Tufts Clinical and Translational Science Institute • Duke University — UL1TR002553: Duke Clinical and Translational Science Institute • Virginia Commonwealth University — UL1TR002649: C. Kenneth and Dianne Wright Center for Clinical and Translational Research • The Ohio State University — UL1TR002733: Center for Clinical and Translational Science • The University of Miami Leonard M. Miller School of Medicine — UL1TR002736: University of Miami Clinical and Translational Science Institute • University of Virginia — UL1TR003015: iTHRIV Integrated Translational health Research Institute of Virginia • Carilion Clinic — UL1TR003015: iTHRIV Integrated Translational health Research Institute of Virginia • University of Alabama at Birmingham — UL1TR003096: Center for Clinical and Translational Science • Johns Hopkins University — UL1TR003098: Johns Hopkins Institute for Clinical and Translational Research • University of Arkansas for Medical Sciences — UL1TR003107: UAMS Translational Research Institute • Nemours — U54GM104941: Delaware CTR ACCEL Program • University Medical Center New Orleans — U54GM104940: Louisiana Clinical and Translational Science (LA CaTS) Center • University of Colorado Denver, Anschutz Medical Campus — UL1TR002535: Colorado Clinical and Translational Sciences Institute • Mayo Clinic Rochester — UL1TR002377: Mayo Clinic Center for Clinical and Translational Science (CCaTS) • Tulane University — UL1TR003096: Center for Clinical and Translational Science • Loyola University Medical Center — UL1TR002389: The Institute for Translational Medicine (ITM) • Advocate Health Care Network — UL1TR002389: The Institute for Translational Medicine (ITM) • OCHIN — INV-018455: Bill and Melinda Gates Foundation grant to Sage Bionetworks • The University of Texas Health Science Center at Houston — UL1TR003167: Center for Clinical and Translational Sciences (CCTS) • NorthShore University HealthSystem UL1TR002389: The Institute for Translational Medicine (ITM) • Weill Medical College of Cornell University UL1TR002384: Weill Cornell Medicine Clinical and Translational Science Center • Montefiore Medical Center — UL1TR002556: Institute for Clinical and Translational Research at Einstein and Montefiore • Medical College of Wisconsin — UL1TR001436: Clinical and Translational Science Institute of Southeast Wisconsin • George Washington University — UL1TR001876: Clinical and Translational Science Institute at Children’s National (CTSA-CN) • Regenstrief Institute — UL1TR002529: Indiana Clinical and Translational Science Institute • Cincinnati Children’s Hospital Medical Center — UL1TR001425: Center for Clinical and Translational Science and Training • Boston University Medical Campus — UL1TR001430: Boston University Clinical and Translational Science Institute • The State University of New York at Buffalo — UL1TR001412: Clinical and Translational Science Institute • Brown University — U54GM115677: Advance Clinical Translational Research (Advance-CTR) • Rutgers, The State University of New Jersey — UL1TR003017: New Jersey Alliance for Clinical and Translational Science • Loyola University Chicago — UL1TR002389: The Institute for Translational Medicine (ITM) • New York University — UL1TR001445: Langone Health’s Clinical and Translational Science Institute • University of Kansas Medical Center — UL1TR002366: Frontiers: University of Kansas Clinical and Translational Science Institute • Massachusetts General Brigham — UL1TR002541: Harvard Catalyst • University of California, Irvine — UL1TR001414: The UC Irvine Institute for Clinical and Translational Science (ICTS) • University of California, San Diego — UL1TR001442: Altman Clinical and Translational Research Institute • University of California, Davis — UL1TR001860: UCDavis Health Clinical and Translational Science Center • University of California, San Francisco — UL1TR001872: UCSF Clinical and Translational Science Institute • University of California, Los Angeles — UL1TR001881: UCLA Clinical Translational Science Institute • University of Vermont — U54GM115516: Northern New England Clinical & Translational Research (NNE-CTR) Network

#### Additional Data Partners Who Have Signed a DTA and Whose Data Release is Pending

The Scripps Research Institute — UL1TR002550: Scripps Research Translational Institute • University of Texas Health Science Center at San Antonio — UL1TR002645: Institute for Integration of Medicine and Science • Yale New Haven Hospital — UL1TR001863: Yale Center for Clinical Investigation • Emory University UL1TR002378: Georgia Clinical and Translational Science Alliance • University of New Mexico Health Sciences Center — UL1TR001449: University of New Mexico Clinical and Translational Science Center • Stanford University — UL1TR003142: Spectrum: The Stanford Center for Clinical and Translational Research and Education • Aurora Health Care — UL1TR002373: Wisconsin Network For Health Research • Children’s Hospital of Philadelphia — UL1TR001878: Institute for Translational Medicine and Therapeutics • Icahn School of Medicine at Mount Sinai — UL1TR001433: ConduITS Institute for Translational Sciences • Ochsner Medical Center — U54GM104940: Louisiana Clinical and Translational Science (LA CaTS) Center • HonorHealth — None (Voluntary) • Arkansas Children’s Hospital — UL1TR003107: UAMS Translational Research Institute

### Authors’ contributions

- data curation, integration, and quality assurance: ERP, AB, KK, EH, JL, RM, CGC
- clinical subject matter expertise: JMB, RW, TDB, HD
- statistical analysis: ERP, CMB, AB, AG
- data visualization: CMB, AB, JAM, AG
- project management: LK
- governance/regulatory oversight: MH, JAM, CGC
- manuscript drafting: ERP, CMB, JMC, AB, HD, AG, EH, LK, KK, JL, JAM, RW, TDB, RM, CGC, MH
- critical revision of the manuscript for important intellectual content: ERP, CMB, JMC, AB, HD, AG, EH, LK, KK, JL, JAM, RW, TDB, RM, CGC, MH

## Acknowledgments

This research was possible because of the patients whose information is included within the data from participating organizations (covid.cd2h.org/dtas) and the organizations and scientists (covid.cd2h.org/duas) who have contributed to the on-going development of this community resource[22].

We thank Daniel Brannock for his contributions to Figure 1.

## Notes

### Competing Interest Statement

AT Girvin is an employee of Palantir Technologies. MA Haendel and JA McMurry are co-founders of Pryzm Health.

### Author Declarations

The IRBs of the University of North Carolina at Chapel Hill, University of Tennessee Health Science Center, University of Rochester, Northeastern University, University of Virginia, University of Colorado, Stony Brook University, and Johns Hopkins University gave ethical approval for this work.

### Summary of Updates

Analysis was updated with significantly more U09.9 patients.

