## Supplemental Materials for "Coding Long COVID: Characterizing a new disease through an ICD-10 lens"

### Supplemental Material

#### Supplemental Methods

##### Community Detection in Diagnosis Analysis

To determine conditions and symptoms that were more likely to co-occur with each other in our sample cohort than at random, we conducted a network analysis of a weighted network with nodes representing individual diagnoses, edges between nodes representing co-occurrence, and edge weights corresponding to the count of patients with both conditions. We employed three community detection algorithms: the Girvan-Newman algorithm, the Walktrap algorithm, and the Louvain algorithm. The Girvin-Newman algorithm is a hierarchical approach based on edge betweenness. Edges with high betweenness usually bridge densely connected clusters, and this algorithm detects and deletes edges with high betweenness in order to detect latent structures, assigns communities accordingly, and iteratively partitions within those communities. The Walktrap algorithm is based on the computation of a transition matrix with each element denoting the probability of one node traversing to another, and then simulating “random walks” for an n number of steps in an iterative process. Nodes are assigned to a community based on the community assignment of their neighbors within the given n distance. The Louvain algorithm iteratively uses modularity to optimize partitions, by first determining initial assignments by exchanging nodes between communities until the optimal modularity is reached, and repeating this iterative process treating the produced communities as nodes, with ties between communities. In the case of our co-occurrence network, the Louvain algorithm outperformed the Walktrap and Girvin-Newman algorithms. The Girvin-Newman algorithm may have been penalized for its choice of betweenness as a selection criterion, due to the presence of multiple edges connecting communities in this dense network. The Walktrap algorithm operates bottom-up, and is not as vulnerable to initial hierarchical partitioning that the Girvin-Newman algorithm is, but tended to categorize small communities that were connected to a central cluster via weaker links, as separate communities, effectively producing smaller, less well-separated clusters. The Louvain algorithm grouped these smaller fragmented clusters along with nearby large clusters, both improving modularity and interpretability of our final communities.

##### Network Stability

We assessed the stability of the co-occurrence network using the quadratic assignment procedure (QAP). The QAP is a resampling-based method (similar to the bootstrap) that measures the correlation between two network matrices and calculates standard errors for associations. For our analysis, we use QAP to provide a statistical assessment of network similarity between the full network and a re-sampled subset of networks (75%, 50%, 25%, and 5% subsamples of patients in the original network). The resulting subsampled network correlation values against the full network were: 0.73 (5% network against the full network), 0.80 (25% network against the full network), 0.81 (50% network against the full network), and 0.81 (75% network against the full network). The results of the QAP analysis were statistically significant at an $\alpha = 0.05$, indicating high network stability.

##### Age-Stratified Condition Co-occurrence Networks

For further subgroup analyses of our co-occurrence network, we presented clusters detected within age-stratified condition networks (age groups <21, 21-45, 46-55, 56-65, 65+; see **Figure 2** for results). In order to determine whether the differences in communities detected across age-stratified cohorts may be attributed to a true difference in prevalence for selected conditions across cohorts, we conducted a Pearson’s chi-square test of independence. Specifically, we examined the statistical significance ($\alpha = 0.05$) of the difference across age-groups in the prevalence for each condition that is prevalent in at least one age cohort (a total of 52 conditions). Although the observed value for the age-stratified prevalence of a few conditions was < 5, the expected value in each cell was > 5 across all conditions, allowing us to use the chi-square test instead of an exact test of independence that accounts for issues associated with small cell sizes. Of the 52 conditions present in all cohorts, the difference in prevalence was statistically significant for all 52 conditions ($p < 0.05$).

### Supplemental Figure 1

**Common conditions among patients with a U09.9 code.** Conditions shown occur within 60 days after a patient's U09.9 diagnosis. Conditions associated with fewer than 20 patients or less than 1% of the age-stratified cohort size are not shown.

*Figure starts on next page.*

| Condition | < 21 years of age (Total = 1490 Patients) % Patients (N) | 21 - 45 years of age (Total = 7263 Patients) % Patients (N) | 46 - 65 years of age (Total = 8600 Patients) % Patients (N) | 66+ years of age (Total = 3719 Patients) % Patients (N) |
|---|---|---|---|---|
| Dyspnea | 18.2% (271) | 32.3% (2348) | 36.5% (3138) | 33.6% (1248) |
| Essential hypertension | - | 10.2% (744) | 25.9% (2231) | 38.3% (1423) |
| Fatigue | 15.7% (234) | 18.7% (1355) | 18.5% (1588) | 14.6% (543) |
| Cough | 13.8% (205) | 15.9% (1152) | 16.9% (1456) | 14.8% (550) |
| Chest pain | 12.2% (182) | 15.3% (1108) | 12.2% (1047) | 7.3% (271) |
| Anxiety disorder | 7.1% (106) | 15.1% (1095) | 11.8% (1017) | 7.8% (291) |
| Gastroesophageal reflux disease w/o esophagitis | 2.9% (43) | 7.0% (506) | 10.8% (927) | 14.0% (521) |
| Hyperlipidemia | - | 3.3% (237) | 10.8% (925) | 20.2% (750) |
| Headache | 10.1% (151) | 9.6% (697) | 8.0% (687) | 4.5% (169) |
| Chronic cough | 7.2% (108) | 7.5% (547) | 7.8% (671) | 7.7% (286) |
| Chronic pain | 4.0% (60) | 7.5% (546) | 9.1% (783) | 9.0% (335) |
| Finding related to attentiveness | 3.2% (48) | 10.3% (748) | 10.3% (884) | 5.3% (198) |
| Obstructive sleep apnea syndrome | 1.3% (20) | 5.0% (363) | 10.7% (919) | 11.6% (431) |
| Type 2 diabetes mellitus w/o complication | - | 3.0% (220) | 10.3% (883) | 14.0% (520) |
| Chronic fatigue syndrome | 4.6% (69) | 7.3% (529) | 7.3% (629) | 7.3% (271) |
| Depressive disorder | 2.9% (43) | 8.0% (580) | 8.1% (693) | 7.0% (260) |
| Dizziness and giddiness | 6.4% (95) | 7.2% (526) | 6.1% (523) | 5.8% (216) |
| Palpitations | 4.8% (72) | 9.8% (710) | 6.1% (523) | 3.1% (117) |
| Obesity | 2.3% (35) | 6.3% (457) | 7.4% (637) | 6.4% (239) |
| Tachycardia | 5.6% (84) | 7.8% (568) | 5.2% (447) | 3.2% (119) |
| Uncomplicated asthma | 4.0% (60) | 7.0% (507) | 6.2% (536) | 4.5% (168) |
| Malaise | 3.6% (53) | 4.6% (331) | 5.9% (511) | 7.5% (278) |
| Abnormal findings on diagnostic imaging of lung | - | 2.9% (211) | 6.3% (543) | 9.0% (334) |
| Hypothyroidism | - | 2.9% (209) | 5.1% (442) | 9.1% (338) |
| Sensory disorder of smell and/or taste | 4.4% (66) | 5.2% (378) | 3.9% (335) | 3.5% (129) |
| Atherosclerosis of coronary artery w/o angina pectoris | - | - | 3.9% (337) | 12.0% (447) |
| Type 2 diabetes mellitus | - | 1.6% (116) | 5.6% (481) | 9.2% (344) |
| Morbid obesity | - | 5.9% (427) | 6.6% (566) | 3.8% (142) |
| Vitamin D deficiency | 1.6% (24) | 4.3% (312) | 5.0% (432) | 5.3% (197) |
| Viral disease | 5.4% (81) | 3.0% (217) | 3.2% (277) | 3.4% (126) |
| Pneumonia caused by SARS-CoV-2 | - | 2.6% (188) | 5.1% (437) | 6.9% (255) |
| Generalized anxiety disorder | 2.7% (40) | 5.3% (388) | 3.7% (316) | 2.3% (85) |
| Blood chemistry abnormal | 2.1% (31) | 3.4% (247) | 3.7% (317) | 4.5% (169) |
| Hypoxemia | - | 2.0% (144) | 4.7% (407) | 6.9% (256) |
| Electrocardiogram abnormal | 3.0% (45) | 2.5% (178) | 3.2% (272) | 4.8% (179) |
| Fever | 7.7% (114) | 2.0% (144) | 2.0% (175) | 1.7% (65) |
| Muscle pain | 3.2% (47) | 4.0% (292) | 3.9% (332) | 2.3% (87) |
| Abdominal pain | 5.7% (85) | 3.0% (221) | 2.4% (204) | 2.2% (83) |
| Mixed hyperlipidemia | - | 1.1% (78) | 4.2% (360) | 7.9% (292) |
| Asthenia | - | 2.4% (177) | 3.5% (302) | 6.9% (256) |
| Anemia | 1.5% (23) | 1.9% (140) | 2.8% (238) | 6.5% (241) |
| Abnormal breathing | 2.8% (42) | 2.8% (205) | 3.8% (331) | 3.1% (117) |
| Chronic obstructive lung disease | - | - | 3.5% (300) | 8.3% (308) |
| Nausea | 3.7% (55) | 3.7% (270) | 2.7% (233) | 2.0% (76) |
| Joint pain | 2.8% (41) | 3.7% (266) | 3.7% (321) | 1.7% (65) |
| Post-viral disorder | 6.1% (91) | 2.5% (183) | 1.9% (161) | 1.3% (50) |
| Insomnia | - | 3.5% (254) | 3.8% (326) | 4.5% (169) |
| Diarrhea | 3.0% (45) | 3.1% (223) | 3.0% (259) | 2.6% (98) |
| Low back pain | - | 3.2% (234) | 4.0% (346) | 4.4% (162) |
| Constipation | 3.8% (57) | 1.8% (133) | 2.1% (180) | 3.7% (137) |
| Allergic rhinitis | 3.6% (53) | 2.7% (199) | 2.7% (228) | 2.2% (83) |
| Sexually active | - | 5.1% (373) | 3.3% (282) | 2.6% (96) |
| Congestive heart failure | - | - | 2.5% (213) | 7.7% (288) |
| Pneumonia | - | 1.6% (115) | 3.1% (270) | 5.3% (197) |
| Nasal congestion | 2.4% (36) | 2.8% (204) | 2.6% (222) | 2.0% (74) |
| Acute pharyngitis | 4.6% (68) | 2.6% (191) | 1.6% (134) | - |
| Disorder of lung | - | 1.7% (124) | 3.7% (316) | 4.2% (157) |
| Acute upper respiratory infection | 3.6% (53) | 2.5% (184) | 1.7% (143) | 1.7% (62) |
| Cardiac arrhythmia | 3.2% (47) | 2.8% (201) | 1.6% (138) | 1.7% (64) |
| Atrial fibrillation | - | - | 1.6% (138) | 7.2% (268) |
| Hyperglycemia due to type 2 diabetes mellitus | - | 1.0% (74) | 3.5% (300) | 4.1% (154) |
| Dependence on supplemental oxygen | - | 1.0% (73) | 3.2% (271) | 4.5% (167) |
| Acute hypoxemic respiratory failure | - | 1.1% (79) | 2.9% (253) | 4.3% (161) |
| Pure hypercholesterolemia | - | - | 2.7% (228) | 4.8% (178) |
| Sexually active with men | - | 3.6% (260) | 2.4% (207) | 2.2% (80) |
| Backache | 1.3% (20) | 1.7% (124) | 2.5% (212) | 2.4% (90) |
| Uncomplicated moderate persistent asthma | 2.4% (36) | 2.1% (154) | 1.9% (162) | 1.5% (56) |
| Mild intermittent asthma | 2.4% (36) | 2.4% (173) | 1.6% (141) | 1.5% (55) |
| Nicotine dependence | - | 2.7% (199) | 3.2% (271) | 1.9% (71) |
| Prediabetes | - | 1.4% (103) | 3.2% (275) | 3.1% (117) |
| Nausea and vomiting | 2.9% (43) | 2.0% (146) | 1.5% (132) | 1.3% (47) |
| Neck pain | - | 2.5% (185) | 2.7% (235) | 2.4% (88) |
| Postoperative state | - | 1.4% (99) | 2.4% (205) | 3.9% (145) |

| | | | | |
|---|---|---|---|---|
| Disorder of body system | 1.5% (23) | 1.7% (125) | 1.9% (164) | 2.3% (84) |
| Osteoarthritis | - | - | 2.5% (212) | 4.4% (162) |
| Interstitial lung disease | - | - | 2.6% (224) | 4.1% (154) |
| Disorder of nasal cavity | 1.5% (23) | 2.2% (158) | 1.9% (164) | 1.7% (65) |
| Loss of consciousness | 2.8% (42) | 1.6% (115) | 1.3% (114) | 1.6% (60) |
| Pulmonary embolism | - | 1.5% (107) | 2.7% (236) | 3.1% (114) |
| Paroxysmal atrial fibrillation | - | - | 1.3% (111) | 5.8% (214) |
| Sleep disorder | 1.7% (26) | 2.1% (153) | 1.9% (165) | 1.3% (47) |
| Amnesia | - | 1.8% (131) | 2.4% (203) | 2.9% (106) |
| Shoulder joint pain | - | 1.4% (103) | 2.7% (235) | 2.9% (106) |
| Paresthesia | - | 2.7% (195) | 2.8% (239) | 1.5% (56) |
| Eruption | 2.6% (39) | 1.6% (115) | 1.5% (126) | 1.3% (48) |
| Postviral fatigue syndrome | 1.5% (23) | 2.2% (162) | 1.8% (158) | 1.3% (49) |
| General finding of observation of patient | 1.9% (29) | 2.0% (143) | 1.6% (138) | 1.4% (51) |
| Iron deficiency anemia | - | 2.0% (145) | 1.9% (162) | 3.0% (111) |
| Wheezing | 2.2% (33) | 1.7% (124) | 1.5% (125) | 1.5% (54) |
| Chronic hypoxemic respiratory failure | - | - | 2.6% (220) | 3.4% (128) |
| Localized edema | - | - | 1.8% (159) | 3.7% (137) |
| Migraine | - | 3.1% (222) | 2.5% (214) | - |
| Heart failure | - | - | 1.6% (139) | 4.3% (161) |
| Reduced mobility | - | 1.6% (118) | 1.9% (160) | 3.0% (111) |
| Polyneuropathy | - | 1.3% (94) | 2.2% (188) | 2.9% (109) |
| Chronic kidney disease due to hypertension | - | - | 1.2% (107) | 5.2% (192) |
| Fibromyalgia | - | 1.8% (132) | 2.6% (221) | 2.0% (75) |
| Osteoarthritis of knee | - | - | 2.5% (216) | 3.8% (141) |
| Major depression, single episode | - | 1.8% (134) | 2.5% (211) | 1.8% (68) |
| Dependence on enabling machine or device | - | - | 2.4% (206) | 2.8% (105) |
| Migraine w/o aura | 1.8% (27) | 2.2% (163) | 1.4% (117) | - |
| Urinary tract infectious disease | - | - | 1.4% (120) | 3.6% (135) |
| Cardiomegaly | - | 1.1% (77) | 1.8% (151) | 2.9% (109) |
| Sleep apnea | - | 1.3% (98) | 2.3% (200) | 2.1% (77) |
| Chronic kidney disease stage 3 | - | - | 1.1% (94) | 4.6% (172) |
| Chronic kidney disease due to type 2 diabetes mellitus | - | - | 1.2% (101) | 4.5% (168) |
| Chronic sinusitis | - | 2.0% (146) | 1.9% (166) | 1.7% (65) |
| Pain | 1.3% (20) | 1.5% (107) | 1.5% (128) | 1.4% (51) |
| Acute renal failure syndrome | - | - | 1.6% (141) | 4.0% (150) |
| Dysphagia | - | 1.2% (88) | 1.8% (154) | 2.6% (96) |
| Vitamin B deficiency | - | 1.4% (102) | 1.6% (135) | 2.6% (97) |
| Hypokalemia | - | - | 1.7% (147) | 3.1% (115) |
| Solitary nodule of lung | - | - | 1.8% (152) | 2.9% (106) |
| Bradycardia | 1.5% (23) | - | 1.1% (91) | 2.1% (77) |
| Currently not sexually active | - | 1.2% (86) | 1.6% (137) | 2.7% (102) |
| Pain in right knee | - | 1.2% (90) | 1.9% (160) | 2.4% (88) |
| Dehydration | 2.3% (35) | - | - | 1.8% (68) |
| Asthma | 1.8% (27) | 1.5% (106) | 1.1% (97) | - |
| Anesthesia of skin | - | 2.2% (157) | 2.1% (184) | - |
| Moderate recurrent major depression | - | 2.0% (142) | 1.7% (148) | 1.3% (49) |
| Pain in left knee | - | - | 1.8% (156) | 2.2% (82) |
| Nervous system symptoms | - | 1.7% (123) | 1.7% (146) | 1.6% (58) |
| Osteoporosis | - | - | - | 4.1% (152) |
| Uncomplicated mild persistent asthma | 2.0% (30) | 1.2% (89) | - | - |
| Disorder of bone | - | - | 1.3% (108) | 3.6% (135) |
| Hypertensive heart failure | - | - | 1.3% (110) | 3.6% (134) |
| Acquired absence of organ | - | - | 1.6% (134) | 2.4% (88) |
| Fibrosis of lung | - | - | 1.7% (148) | 3.1% (114) |
| Snoring | - | 1.9% (135) | 1.9% (165) | - |
| Chronic diastolic heart failure | - | - | 1.0% (84) | 3.7% (137) |
| Hip pain | - | - | 1.7% (143) | 2.0% (74) |
| Dysuria | - | 1.3% (94) | 1.3% (110) | 1.9% (69) |
| Musculoskeletal finding | - | 1.3% (95) | 1.6% (134) | 1.6% (58) |
| Disorder of soft tissue | - | - | 1.6% (137) | 1.9% (69) |
| Inflammatory dermatosis | 1.7% (26) | - | - | - |
| Chronic kidney disease | - | - | 1.1% (97) | 3.2% (118) |
| Radiology result abnormal | - | - | 1.3% (110) | 1.9% (69) |
| Non-scarring alopecia | - | 1.6% (119) | 1.6% (134) | - |
| Acquired hypothyroidism | - | - | 1.1% (93) | 2.2% (83) |
| Loss of sense of smell | - | 1.5% (112) | 1.4% (118) | 1.0% (39) |
| Gastroesophageal reflux disease | - | 1.0% (73) | 1.2% (102) | 1.7% (65) |
| Polyneuropathy due to type 2 diabetes mellitus | - | - | 1.2% (107) | 2.7% (100) |
| Finding of frequency of urination | - | - | 1.1% (97) | 2.0% (75) |
| Immunodeficiency disorder | - | - | 1.5% (128) | 1.6% (61) |
| Chronic pain syndrome | - | - | 1.4% (119) | 1.6% (58) |
| Taste sense altered | - | 1.4% (104) | 1.4% (119) | 1.0% (38) |
| Finding relating to drug misuse behavior | - | 1.0% (76) | 1.3% (115) | 1.5% (54) |
| Bronchitis | - | 1.4% (104) | 1.1% (98) | 1.3% (47) |
| Steatosis of liver | - | 1.2% (84) | 1.8% (151) | - |
| Dysphonia | - | 1.0% (73) | 1.8% (151) | 1.0% (39) |
| Epigastric pain | - | 1.6% (118) | 1.2% (103) | - |
| Hypo-osmolality and or hyponatremia | - | - | 1.1% (91) | 2.7% (100) |

| | | | | |
|---|---|---|---|---|
| Pulmonary emphysema | - | - | 1.1% (91) | 2.6% (98) |
| Edema | - | - | - | 2.0% (75) |
| Chronic rhinitis | - | 1.1% (82) | 1.2% (103) | 1.3% (49) |
| Posttraumatic stress disorder | - | 2.0% (148) | 1.6% (138) | - |
| Sexually active with women | - | 1.3% (93) | - | 1.5% (55) |
| Muscle weakness | - | - | 1.3% (109) | 1.3% (50) |
| Pulmonary hypertension | - | - | 1.1% (97) | 2.4% (91) |
| Visual disturbance | - | 1.3% (95) | 1.3% (116) | - |
| Leukocytosis | - | - | 1.0% (82) | 1.8% (68) |
| Anxiety state | - | 1.5% (108) | 1.3% (109) | - |
| Hyperglycemia | - | - | 1.3% (113) | 1.6% (59) |
| Ventricular premature complex | - | - | 1.0% (86) | 1.7% (62) |
| Lumbar radiculopathy | - | - | 1.4% (117) | 1.5% (54) |
| Attention deficit hyperactivity disorder | 1.9% (29) | 1.5% (107) | - | - |
| Spasm | - | - | 1.3% (110) | 1.2% (43) |
| Disorder of nervous system due to type 2 diabetes mellitus | - | - | 1.3% (110) | 2.0% (76) |
| Diaphragmatic hernia | - | - | 1.1% (95) | 2.2% (82) |
| Pain in right lower limb | - | - | 1.3% (110) | 1.0% (39) |
| Pain in left lower limb | - | - | 1.2% (103) | 1.2% (46) |
| Lumbago with sciatica | - | - | 1.1% (94) | 1.3% (47) |
| Pleural effusion | - | - | - | 2.4% (88) |
| Localized enlarged lymph nodes | - | - | 1.0% (83) | 1.6% (61) |
| Benign prostatic hyperplasia | - | - | - | 2.6% (97) |
| Tremor | - | 1.1% (77) | - | 1.2% (46) |
| Complication due to diabetes mellitus | - | - | 1.5% (129) | 1.7% (63) |
| Generalized abdominal pain | 1.6% (24) | - | - | - |
| Posterior rhinorrhea | - | 1.0% (73) | 1.0% (88) | 1.1% (40) |
| Hypersomnia | - | 1.2% (88) | 1.2% (100) | - |
| Supraventricular tachycardia | - | - | - | 1.5% (57) |
| Vomiting | 2.6% (38) | - | - | - |
| Pulmonary function studies abnormal | - | - | 1.3% (113) | - |
| Bronchiectasis | - | - | 1.0% (89) | 2.0% (75) |
| Cardiomyopathy | - | - | 1.0% (82) | 1.5% (57) |
| Chronic kidney disease stage 3A | - | - | - | 2.5% (92) |
| Benign prostatic hypertrophy with outflow obstruction | - | - | - | 2.4% (88) |
| Old myocardial infarction | - | - | - | 2.2% (82) |
| Low blood pressure | - | - | - | 2.2% (83) |
| Acute sinusitis | - | 1.1% (83) | - | - |
| Tear film insufficiency | - | - | 1.2% (101) | 1.8% (66) |
| Chronic congestive heart failure | - | - | - | 2.3% (87) |
| Urogenital finding | - | - | - | 1.3% (50) |
| Kidney stone | - | - | - | 1.1% (42) |
| Chronic systolic heart failure | - | - | - | 2.3% (85) |
| Neurosis | - | 1.7% (123) | 1.2% (99) | - |
| Multisystem inflammatory syndrome (MIS) | 2.8% (42) | - | - | - |
| Seasonal allergic rhinitis | - | 1.1% (83) | - | - |
| Loss of appetite | - | - | - | 1.5% (57) |
| Disorder of intestine | - | - | 1.0% (84) | 1.1% (40) |
| Coronary artery graft present | - | - | - | 2.2% (81) |
| Peripheral vascular disease | - | - | - | 2.1% (77) |
| Disorder of muscle | - | - | 1.1% (91) | - |
| Rheumatoid arthritis | - | - | 1.2% (103) | 1.5% (54) |
| Non-toxic uninodular goiter | - | - | 1.0% (84) | 1.1% (41) |
| Degeneration of lumbar intervertebral disc | - | - | 1.1% (97) | 1.5% (54) |
| Spinal stenosis of lumbar region | - | - | - | 1.9% (69) |
| Atelectasis | - | - | 1.0% (89) | 1.5% (55) |
| Lumbar spondylosis | - | - | 1.1% (98) | 1.4% (51) |
| Carpal tunnel syndrome | - | - | 1.2% (100) | - |
| Bacterial infectious disease | - | 1.1% (78) | - | - |
| Hemorrhoids | - | - | - | 1.0% (39) |
| Allergic rhinitis due to pollen | - | 1.0% (75) | - | - |
| Hypertensive heart and renal disease with (congestive) heart failure | - | - | - | 2.4% (89) |
| Chronic kidney disease stage 3B | - | - | - | 2.4% (88) |
| Restless legs | - | - | - | 1.5% (54) |
| Hypomagnesemia | - | - | - | 1.6% (58) |
| Gout | - | - | - | 1.7% (62) |
| Respiratory failure | - | - | - | 1.4% (52) |
| Panic disorder w/o agoraphobia | - | 1.4% (103) | - | - |
| Acute exacerbation of chronic obstructive airways disease | - | - | - | 1.5% (56) |
| Menopause present | - | - | - | 1.6% (59) |
| Presbyopia | - | - | 1.1% (92) | 1.2% (44) |
| Senile hyperkeratosis | - | - | - | 1.3% (50) |
| Non-rheumatic mitral valve stenosis with regurgitation | - | - | - | 1.5% (57) |
| Cerebral infarction | - | - | - | 1.5% (55) |
| Disorder of kidney and/or ureter | - | - | - | 1.6% (60) |
| Sepsis | - | - | - | 1.5% (54) |

| | | | | |
|---|---|---|---|---|
| Inflammatory disorder | 2.2% (33) | - | - | - |
| Peripheral circulatory disorder due to type 2 diabetes mellitus | - | - | - | 1.6% (58) |
| Migraine with aura | - | 1.5% (106) | - | - |
| Cognitive communication disorder | - | 1.2% (88) | 1.0% (84) | - |
| Diastolic heart failure | - | - | - | 1.6% (58) |
| Thrombocytopenic disorder | - | - | - | 1.5% (54) |
| Hyperkalemia | - | - | - | 1.5% (57) |
| Systemic disease | 2.1% (32) | - | - | - |
| Acute on chronic hypoxemic respiratory failure | - | - | - | 1.5% (54) |
| Nuclear senile cataract | - | - | - | 1.5% (57) |
| Benign essential hypertension | - | - | - | 1.4% (51) |
| Anemia in chronic kidney disease | - | - | - | 1.5% (57) |
| Post-inflammatory pulmonary fibrosis | - | - | - | 1.2% (43) |
| Allergic disposition | - | 1.3% (94) | - | - |
| Heart disease | - | - | - | 1.4% (52) |
| Acidosis | - | - | - | 1.5% (54) |
| Aortocoronary bypass graft present | - | - | - | 2.0% (75) |
| Centriacinar emphysema | - | - | - | 1.5% (55) |
| Actinic keratosis | - | - | - | 1.4% (51) |
| Embolism from thrombosis of vein of lower extremity | - | - | - | 1.2% (43) |
| Right bundle branch block | - | - | - | 1.4% (52) |
| Sensorineural hearing loss, bilateral | - | - | - | 1.4% (52) |
| Systolic heart failure | - | - | - | 1.3% (50) |
| Localized, primary osteoarthritis of the shoulder region | - | - | - | 1.2% (46) |
| Diverticulosis of large intestine w/o diverticulitis | - | - | - | 1.2% (45) |
| Erectile dysfunction | - | - | - | 1.0% (38) |
| Orthostatic hypotension | - | - | - | 1.2% (45) |
| End-stage renal disease | - | - | - | 1.2% (44) |
| Osteoarthritis of hip | - | - | - | 1.3% (47) |
| Hearing loss | - | - | - | 1.3% (50) |
| Difficulty walking | - | - | - | 1.1% (40) |
| Chronic kidney disease stage 4 | - | - | - | 1.8% (67) |
| Noninflammatory disorder of the vagina | - | 1.2% (88) | - | - |
| Impacted cerumen | - | - | - | 1.2% (43) |
| Primary malignant neoplasm of prostate | - | - | - | 1.7% (62) |
| Urinary incontinence | - | - | - | 1.1% (41) |
| Parosmia | - | 1.0% (75) | - | - |
| Disorder of brain | - | - | - | 1.0% (38) |
| Altered mental status | - | - | - | 1.0% (39) |
| Chronic kidney disease stage 2 | - | - | - | 1.5% (57) |
| Finding related to pregnancy | - | 1.5% (111) | - | - |
| Pain in pelvis | - | 1.4% (105) | - | - |
| Dementia | - | - | - | 1.3% (50) |
| Acute cystitis | - | - | - | 1.3% (49) |
| Disorientated | - | - | - | 1.3% (47) |
| Peripheral venous insufficiency | - | - | - | 1.3% (47) |
| Acute on chronic diastolic heart failure | - | - | - | 1.2% (46) |
| Transplanted heart valve present | - | - | - | 1.2% (46) |
| First degree atrioventricular block | - | - | - | 1.2% (45) |
| Carotid artery obstruction | - | - | - | 1.2% (44) |
| Raised prostate specific antigen | - | - | - | 1.2% (44) |
| Coronary atherosclerosis | - | - | - | 1.2% (43) |
| Dementia associated with another disease | - | - | - | 1.2% (43) |
| Hypertensive heart disease w/o congestive heart failure | - | - | - | 1.2% (43) |
| Non-rheumatic aortic sclerosis | - | - | - | 1.2% (43) |
| Sick sinus syndrome | - | - | - | 1.1% (42) |
| Complication occurring during pregnancy | - | 1.1% (78) | - | - |
| Disorder of pregnancy | - | 1.1% (78) | - | - |
| High risk pregnancy | - | 1.1% (78) | - | - |
| Acne | - | 1.1% (77) | - | - |
| Acute posthemorrhagic anemia | - | - | - | 1.0% (39) |
| Ventricular tachycardia | - | - | - | 1.0% (39) |
| Familial dysautonomia | - | 1.0% (76) | - | - |
| Persistent atrial fibrillation | - | - | - | 1.0% (38) |
| Unsteady when standing | - | - | - | 1.0% (38) |
| Genitourinary tract hemorrhage | - | 1.0% (74) | - | - |

### Supplemental Figure 2

**Common medications among patients with a U09.9 code.** Medications shown occur within 60 days after a patient's U09.9 diagnosis, and do *not* occur prior to the U09.9 (i.e., new medications). Medications are coded using the ATC terminology. Because a single drug can have multiple ATC codes, some medications are counted in more than one category. Category totals represent unique patient - drug pairs, not necessarily unique individuals. Medication classes associated with fewer than 20 patients or less than 0.5% of the age-stratified cohort size are not shown, per N3C download policy. Percentages in each column are shown relative to the total *n* in that column.

When using EHR data, it can be difficult to discern indication from drug records, particularly when drugs are recorded at the ingredient level, and particularly when those ingredients can be used in a wide variety of medications and medication forms. For this reason, the categories of Opthalmologicals; Otologicals; Corticosteroids, Dermatological Preparations; and Blood Substitutes and Perfusion Solutions are artificially inflated. They are included for completeness, but should not be interpreted at face value.

| Medication Group | < 21 years of age (Total = 1490 Patients) | 21 - 45 years of age (Total = 7263 Patients) | 46 - 65 years of age (Total = 8600 Patients) | 66+ years of age (Total = 3719 Patients) |
|---|---|---|---|---|
| | % Patients (N) | % Patients (N) | % Patients (N) | % Patients (N) |
| **Respiratory System Drugs** | | | | |
| Drugs For Obstructive Airway Diseases | 10.3% (153) | 10.4% (755) | 9.0% (773) | 7.0% (262) |
| Nasal Preparations | 3.9% (58) | 3.2% (233) | 3.1% (263) | 2.5% (93) |
| Cough And Cold Preparations | 2.4% (35) | 3.5% (253) | 3.8% (326) | 3.2% (117) |
| Antihistamines For Systemic Use | 1.9% (28) | 1.4% (100) | 1.3% (112) | - |
| Throat Preparations | 1.7% (25) | - | - | - |
| **Alimentary Tract and Metabolism Drugs** | - | - | - | - |
| Antidiarrheals, Intestinal Antiinflammatory/Antiinfective Age | 5.4% (80) | 4.8% (346) | 5.1% (439) | 5.4% (200) |
| Mineral Supplements | 4.4% (65) | 2.0% (144) | 2.8% (243) | 2.9% (108) |
| Drugs For Acid Related Disorders | 3.8% (56) | 2.0% (145) | 2.3% (201) | 2.0% (75) |
| Vitamins | 1.8% (27) | 2.6% (191) | 3.1% (269) | 2.3% (87) |
| Antiemetics And Antinauseants | 2.2% (32) | - | 1.1% (97) | 1.2% (45) |
| Stomatological Preparations | 2.2% (32) | 1.4% (99) | 1.3% (113) | - |
| Drugs For Constipation | - | - | 1.4% (116) | - |
| Drugs Used In Diabetes | - | - | 1.2% (104) | 1.1% (42) |
| **Sensory Organ Drugs** | | | | |
| Ophthalmologicals | 11.7% (174) | 8.6% (626) | 8.2% (706) | 7.5% (279) |
| Otologicals | 7.6% (113) | 4.5% (330) | 4.6% (396) | 3.4% (126) |
| **Cardiovascular System Drugs** | - | - | - | - |
| Cardiac Therapy | 3.9% (58) | 2.0% (146) | 2.7% (235) | 3.6% (132) |
| Diuretics | 4.2% (63) | 2.0% (147) | 3.9% (338) | 3.9% (145) |
| Beta Blocking Agents | 1.8% (27) | 2.4% (176) | 3.6% (310) | 3.4% (125) |
| Vasoprotectives | 3.5% (52) | 1.9% (136) | 2.2% (186) | 2.0% (75) |
| Agents Acting On The Renin-Angiotensin System | - | 1.3% (96) | 3.4% (291) | 3.3% (124) |
| Lipid Modifying Agents | - | 1.0% (73) | 2.4% (202) | 1.9% (70) |
| Calcium Channel Blockers | - | - | - | 1.6% (59) |
| **Dermatologicals** | | | | |
| Corticosteroids, Dermatological Preparations | 8.7% (130) | 6.7% (483) | 5.8% (496) | 3.9% (143) |
| Antipruritics, Incl. Antihistamines, Anesthetics, Etc. | 2.0% (29) | - | 1.1% (97) | 1.0% (38) |
| Other Dermatological Preparations | - | - | - | 1.1% (41) |
| Anti-Acne Preparations | - | 1.0% (73) | - | - |
| **Blood and Blood Forming Organ Drugs** | | | | |
| Blood Substitutes And Perfusion Solutions | 7.6% (113) | 4.6% (333) | 5.2% (445) | 4.5% (169) |
| Antianemic Preparations | - | 1.8% (129) | 1.8% (158) | 1.5% (56) |
| Antithrombotic Agents | 1.7% (25) | - | 1.4% (122) | 1.4% (51) |
| **Nervous System Drugs** | | | | |
| Analgesics | 4.0% (59) | 2.5% (183) | 2.6% (227) | 1.8% (67) |
| Psycholeptics | 1.7% (26) | 1.9% (140) | 2.3% (198) | 2.6% (95) |
| Anesthetics | 2.2% (33) | 1.5% (109) | 1.8% (155) | 1.7% (63) |
| Psychoanaleptics | 2.4% (35) | 1.9% (140) | 1.5% (127) | - |
| **Various Drug Classes in ATC** | | | | |
| Contrast Media | 2.6% (38) | 3.4% (249) | 3.3% (284) | 4.3% (160) |
| Diagnostic Agents | - | - | 1.0% (86) | 1.3% (48) |
| **Antiinfectives for Systemic Use** | | | | |
| Antibacterials For Systemic Use | 4.6% (69) | 4.4% (318) | 5.5% (472) | 5.7% (213) |
| Vaccines | 2.0% (29) | - | - | - |
| **Musculo-Skeletal System Drugs** | | | | |
| Antiinflammatory And Antirheumatic Products | 2.6% (39) | 1.7% (123) | 1.9% (159) | 1.4% (52) |
| Topical Products For Joint And Muscular Pain | 1.4% (21) | - | - | - |
| Drugs For Treatment Of Bone Diseases | - | 1.0% (75) | 1.2% (107) | - |
| **Genitourinary System and Sex Hormones** | | | | |
| Gynecological Antiinfectives And Antiseptics | - | 1.2% (84) | 1.6% (138) | 1.1% (39) |
| Other Gynecologicals | 2.1% (31) | 1.2% (85) | - | - |
| **Other Systemic Hormonal Preparations** | | | | |
| Corticosteroids For Systemic Use | 2.6% (39) | 2.5% (180) | 2.3% (196) | 1.9% (71) |
